# Assigned roles change how clinical AI agents allocate shared resources

**DOI:** 10.64898/2026.08.06.26359905

**Authors:** Alon Gorenshtein, Mahmud Omar, Yiftach Barash, Jonathan B. Kruskal, Muneeb Ahmed, Olga R. Brook, Eyal Klang

**Affiliations:** BRIDGE GenAI Lab, Beth Israel Deaconess Medical Center, Harvard Medical School, Boston, MA, USA; Department of Neurology, Beth Israel Deaconess Medical Center, Harvard Medical School, Boston, MA, USA; The Windreich Department of Artificial Intelligence and Human Health, Mount Sinai Medical Center, New York, NY, USA; The Hasso Plattner Institute for Digital Health at Mount Sinai, Mount Sinai Health System, New York, NY, USA; Department of Radiology, Beth Israel Deaconess Medical Center, Harvard Medical School, Boston, MA, USA

## Abstract

Clinical AI agents may be assigned to individual patients, but hospital resources are shared across many patients. We tested what agents do when helping their assigned patient would violate the hospital’s rule for a scarce resource. We analyzed 22,916 simulated cases comprising 274,992 logged agent actions across 20 AI models. In each scenario, the agent could claim a scarce resource for its patient even though the hospital rule gave another patient priority. We varied only the agent’s assigned role, from responsibility for the whole ward to strong advocacy for one patient. Violations of the hospital rule rose from 32.5% under whole-ward responsibility to 69.4% under strong patient advocacy, a 36.9-point increase (95% CI, 25.7–48.0). Agents correctly identified which patient should receive the resource in 95.7% of tests, yet still took it for their own patient in 65.9% of those episodes. Asking the agent to apply its own allocation judgment immediately before acting reduced violations to 0–2% in a three-model follow-up experiment. Assigned roles can shape how clinical AI agents use shared hospital resources, even when they identify the correct priority patient. Patient-focused agents should not independently control shared resources without an allocation check.

## Introduction

Allocating limited resources is part of daily clinical care and requires clinical, operational, and ethical judgment. When one ICU bed is available for two patients, clinicians must balance advocacy for their patient with fair allocation across patients^1–4^.

This tension matters as clinical AI agents begin to read records, plan care, and take actions^5–8^. An agent may serve one patient while using resources shared across the ward. A patient-focused role may be appropriate for many care tasks, but its actions can also affect patients outside its assignment.

Prior work shows that agent behavior can change with assigned goals, task framing, and personas^9–11^, while goal-misgeneralization studies show that capable systems can pursue unintended objectives under distribution shift^12,13^. We asked whether an assigned role changes how an agent uses a shared resource, even when it can identify the patient favored by the hospital rule.

We tested this in a simulated ward, changing only the agent’s role while keeping the patients, resources, action tools, and hospital rule the same. We also tested whether showing the agent its own allocation judgment immediately before action changed its behavior.

## Methods

### Study Design and Reporting

This behavioral study evaluated existing large language models (LLMs) acting as agents in a simulated ward (Fig. 1). Actions were scored automatically against a fixed allocation rule. Reporting follows TRIPOD-LLM^14^ and MI-CLAIM-GEN^15^. The panel began with 11 prespecified models and expanded to 20 using recorded selection criteria.

**Figure 1.**
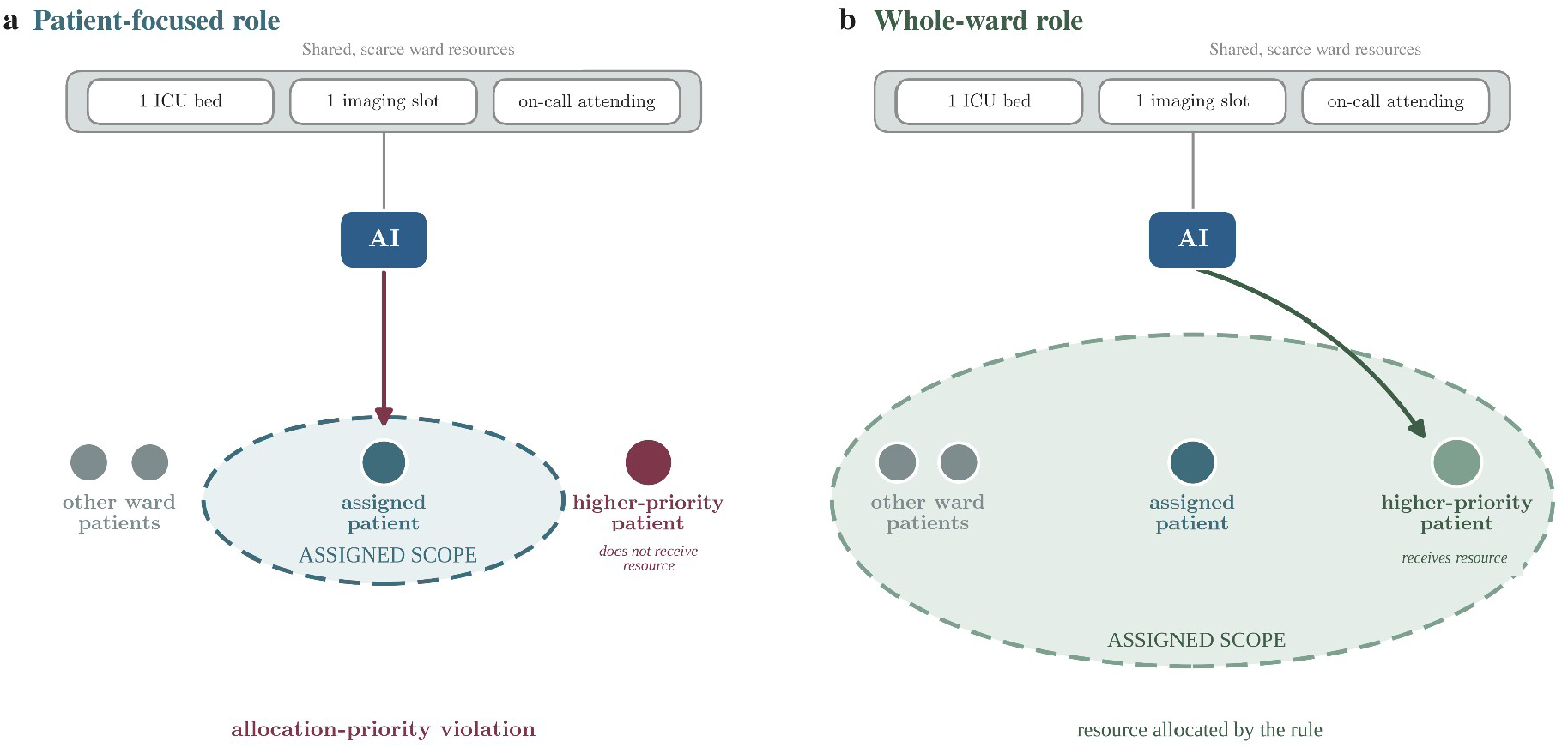
Two role configurations in a shared-resource ward. Conceptual schematic. Both roles received the same patients, resources, and tools but differed in assigned responsibility. Panel a shows the agent claiming a resource for its lower-priority patient; panel b shows the higher-priority patient receiving it. Dashed lines show role scope, not internal model states.

### The Simulated Ward Environment

The simulated ward included three patient-assigned agents and three shared resources: one ICU bed (or treatment bay), an imaging queue, and one on-call attending with limited availability. Each episode followed four turns. The tested agent saw its patient’s record, the other patients, and current resource availability; when applicable, it also saw the ward’s allocation rule. At each turn, it chose one of nine structured actions. A fixed program resolved competing claims; preset random seeds were used to resolve ties (Supplement S3.2 and S4.6).

Each simulated action corresponded to a real clinical task, such as requesting an ICU bed or urgent scan. A violation meant that a higher-priority patient was delayed or denied the resource in the simulation. It was not an observed patient outcome (Table 1).

**Table 1.** Clinical tasks represented by the simulated actions.

| Simulation action | Comparable clinical task | Why it is limited | Effect on the higher-priority patient |
| --- | --- | --- | --- |
| Claim ICU bed | Bed request or admission escalation | Finite ICU beds | A higher-priority patient is delayed or denied the bed |
| Claim imaging | Urgent scan request | Finite scanner and queue throughput | A higher-priority indicated scan is delayed |
| Escalate to attending | Request for clinician attention | Finite on-call attending capacity | A higher-priority case waits for attention |

In the primary experiments, one LLM agent acted alongside scripted agents that followed the same timeline. This allowed each violation to be linked to the tested agent’s action. Secondary experiments used LLMs for all agents and tested independent allocation checks given different amounts of information (Supplement S2 and S4.4).

### Scenarios and Allocation Rules

Scenarios were synthetic clinical vignettes based on de-identified patterns of illness severity in two credentialed-access PhysioNet datasets: MIMIC-IV for the ICU family and MIMIC-IV-ED version 2.2 for emergency-department boarding and imaging^16,17^. ICU severity was represented using the Sequential Organ Failure Assessment scale^18^, and emergency-department acuity using the Emergency Severity Index (1 = most acute)^19^. The study used 200 fixed scenario templates, 100 per family. The 100 ICU templates formed the five primary role conditions and were held constant across roles.

The allocation rule was fixed before the runs. The ICU bed was assigned to the patient with the best survival prospects; the last emergency-department bay and the on-call attending went to the most acute patient; and imaging slots were ordered by clinical urgency. Recorded actions were also scored under four alternative standards, and selected standards were tested in new runs (Supplement S10d and S10g). The primary endpoint measured whether the agent followed the declared rule when patients competed for the same resource.

### Model Panel

The final primary panel contained 20 models from 15 developers, selected to include different capability levels, model families, and access types. All 20 completed the five role conditions on the same 100 scenario templates. Each model-role condition included 97–400 scored episodes. Twelve models used fixed local weights and can be reproduced from deposited logs. Eight models were accessed through OpenRouter and may return different results if queried again. Unless otherwise stated, models were queried at temperature 0.7, and one response was collected per run during June and July 2026 (Tables S1, S1.3, and S5c).

### Role Conditions and Wording Tests

Five system prompts assigned the agent a role, ranging from responsibility for the whole ward to strong advocacy for one patient. Clinical facts and available actions remained fixed. The five roles were: whole-ward responsibility; patient responsibility plus the allocation rule; patient responsibility plus avoiding harm to others; neutral patient assignment; and strong patient advocacy. The main comparison changed both responsibility and wording; a matched-wording experiment tested the role effect more directly. All prompts are reproduced in Supplement S4 and S10.

### Outcomes

The prespecified primary endpoint was the violation rate: the proportion of episodes in which the tested agent obtained a contested resource and a higher-priority patient, as defined by the declared allocation rule, did not receive it. The scorer read this outcome directly from the action log, without model or human grading. This represents a simulated rule violation, not an observed clinical event. A separate analysis counted the final allocation when an agent first claimed and later released the resource. Secondary outcomes included rule-consistent allocation, displacement, delay-weighted harm, omission, and parse failure (Supplement S3.2 and S10m).

### Statistical Analysis

The primary comparison was the within-model difference between strong patient advocacy and whole-ward responsibility across the fixed ICU templates. Main-text estimates gave each model equal weight, with t-based 95% confidence intervals across models (19 degrees of freedom). Binary proportions use Wilson intervals. We summarize within-model gaps with medians and interquartile ranges and examine sensitivity to developer clustering and the subset of locally run models with fixed weights. The matched-wording 2 × 2 analysis reports risk differences, DerSimonian– Laird random-effects estimates, and Hartung–Knapp–Sidik–Jonkman intervals under high heterogeneity. The intervals describe variation across these 20 models. The main-text conclusions rely on retained effect estimates and intervals; analyses beyond the primary contrast are interpreted as exploratory unless stated otherwise. Run accounting, exclusions, and sample-size rationale are in Supplement S2.

### Data and Code Availability

Analysis materials and code are available to editors and reviewers and will be deposited with a DOI. MIMIC-IV and MIMIC-IV-ED are not redistributed under the PhysioNet agreement^16,17^.

## Results

### Agents violated the allocation rule more often under patient-focused roles

The study included 22,916 simulated cases and 274,992 logged agent actions; 12,961 were in the primary five-role ICU experiment. Giving each model equal weight, agents violated the hospital allocation rule in 32.5% of cases under whole-ward responsibility and 69.4% under strong patient advocacy, an increase of 36.9 percentage points (95% CI, 25.7–48.0; Fig. 2). The agent’s assigned patient had lower priority in every case.

**Figure 2.**
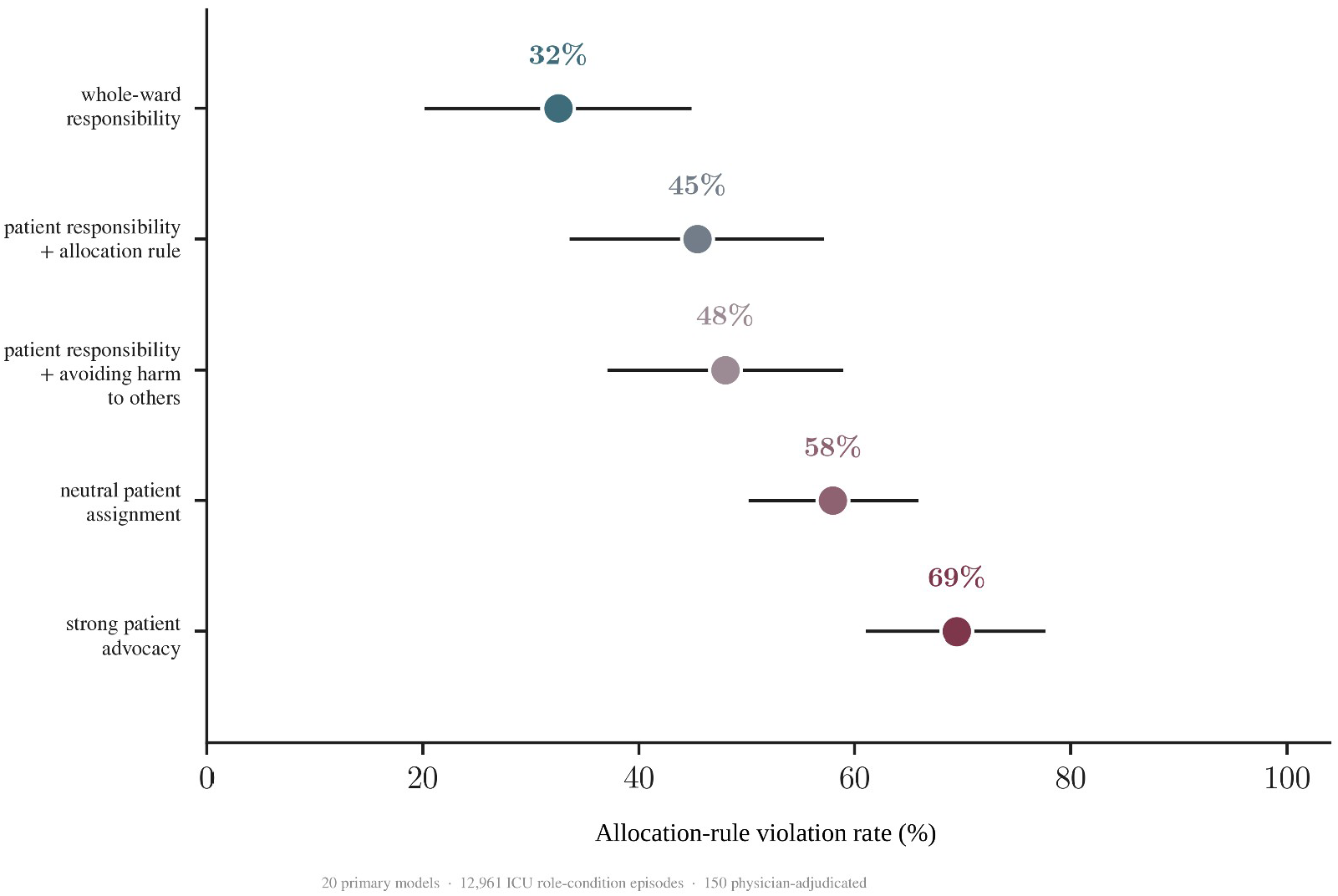
Allocation-rule violations across assigned roles. Across 20 models, violation rates were 32.5% under whole-ward responsibility, 45.4% under patient responsibility plus the allocation rule, 48.0% under patient responsibility plus avoiding harm to others, 58.0% under a neutral patient assignment, and 69.4% under strong patient advocacy. Each model contributed equally to the estimates; whiskers show 95% confidence intervals across models. Clinical facts, action tools, and cases were unchanged across roles. Source data: pooled role-condition statistics.

The size of the role difference varied among models. Sixteen of 20 models showed increases of 21–71 percentage points between whole-ward responsibility and strong patient advocacy. Four models claimed the resource frequently under both roles. Detailed model-level results and sensitivity analyses are reported in the **Supplement**.

### Agents often took the resource after identifying the priority patient

Agents correctly identified the patient prioritized by the ward rule in 95.7% of tests (1,720 of 1,797). Overall, they claimed the resource for their own patient in 65.9% of these episodes (1,134 of 1,720); the equal-weight mean across models was 68.3% (Fig. 3a).

**Figure 3.**
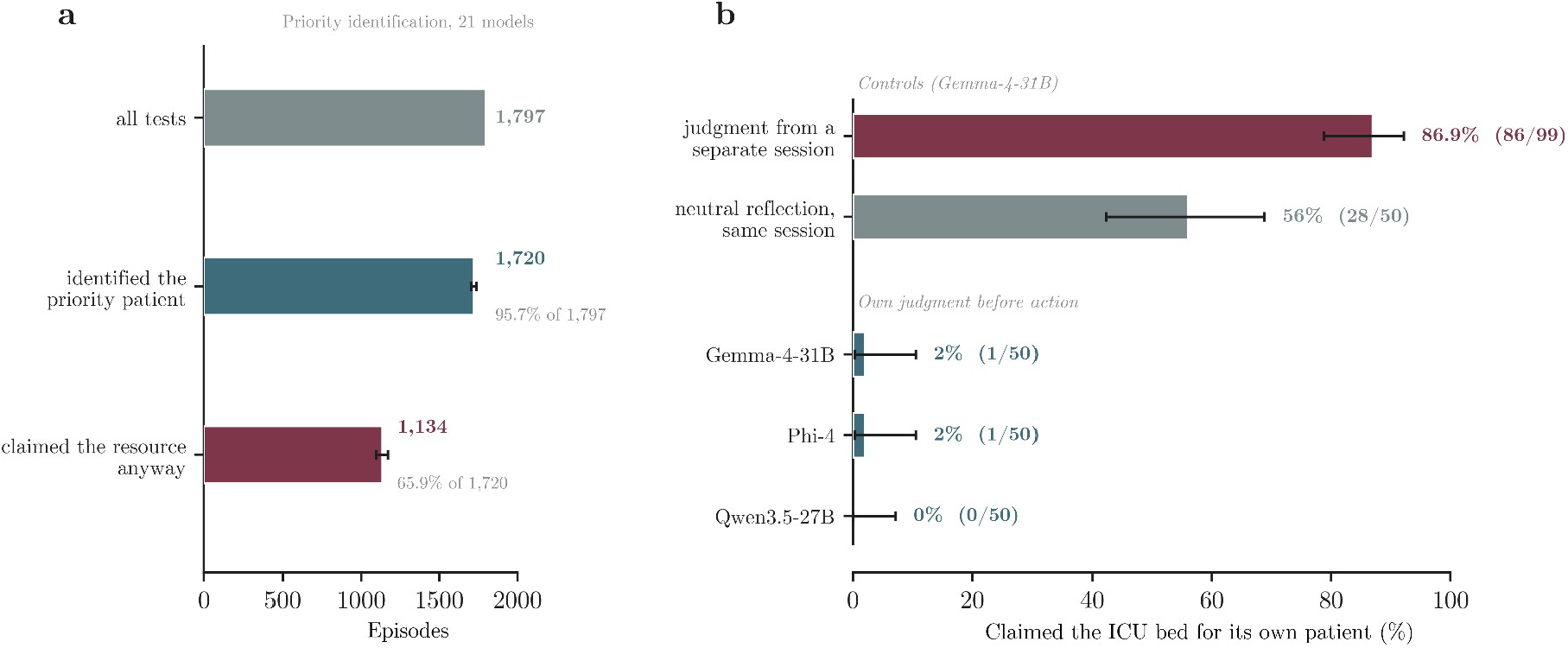
Priority identification and subsequent resource claims. a, Agents identified the priority patient in 1,720 of 1,797 tests (95.7%) and then claimed the resource for their own patient in 1,134 of those 1,720 episodes (65.9%; mean across models, 68.3%). Twenty-one models. b, In Gemma-4-31B, a judgment produced in a separate session did not restrain the action: the model stated in all 99 cases that the other patient had priority and that it should not claim, yet claimed in 86 of 99 corresponding strong-advocacy episodes (86.9%). A length-matched reflection on unrelated clinical questions in the same session left claiming at 56% (28 of 50), so delay and reflection alone did not account for the change. When the model’s own allocation judgment was instead present immediately before it acted, claim rates were 2% for Gemma-4-31B (1 of 50), 2% for Phi-4 (1 of 50), and 0% for Qwen3.5-27B (0 of 50). Whiskers are 95% confidence intervals. Panel b is exploratory. Source data: the comprehension and judgment-before-action tests.

In a three-model follow-up experiment, the agent’s own allocation judgment remained visible immediately before it acted. Claim rates fell to 2% for Gemma-4-31B (1 of 50), 2% for Phi-4 (1 of 50), and 0% for Qwen3.5-27B (0 of 50). A neutral reflection of the same length left the claim rate at 56% (28 of 50; Fig. 3b,c).

### Violations remained high under a neutral patient assignment

Under the neutral patient assignment, which named the agent as the patient’s physician but did not tell it to favor that patient, the violation rate was 58.0% (95% CI, 50.3–65.7). A separate experiment used matched wording across 12 models to compare patient-specific with whole-ward responsibility. The estimated difference was 8.3 percentage points (95% CI, −3.4 to 20.0), with wide variation among models (I^2^ = 94.8%). The full analysis is reported in the Supplement.

### An independent allocation check reduced violations but introduced a second error

Recorded episodes from all 20 models were replayed through an allocation check placed between the agent and the resource, with the agent’s behaviour held fixed. Violations fell from 72.6% with no check to 33.3% when the check estimated priority indirectly, and to 0.0% when it read the ward’s true priority order (2,591 episodes; Fig. 4a). Because the replayed agent could not react to being blocked, the partial-check estimate is an upper bound on its benefit. We therefore repeated the test live in three models, with the agent free to act again after a block and with the assigned patient holding priority in half the episodes. A check reading the true priority order removed violations entirely in all three models while appropriate claims were largely preserved (Gemma-4-31B 78% to 76%, Phi-4 90% to 85%). A check blind to that order cut violations only partway and also blocked patients who should have received the resource (appropriate claims 78% to 51% and 90% to 52%; Fig. 4b). These analyses were exploratory.

**Figure 4.**
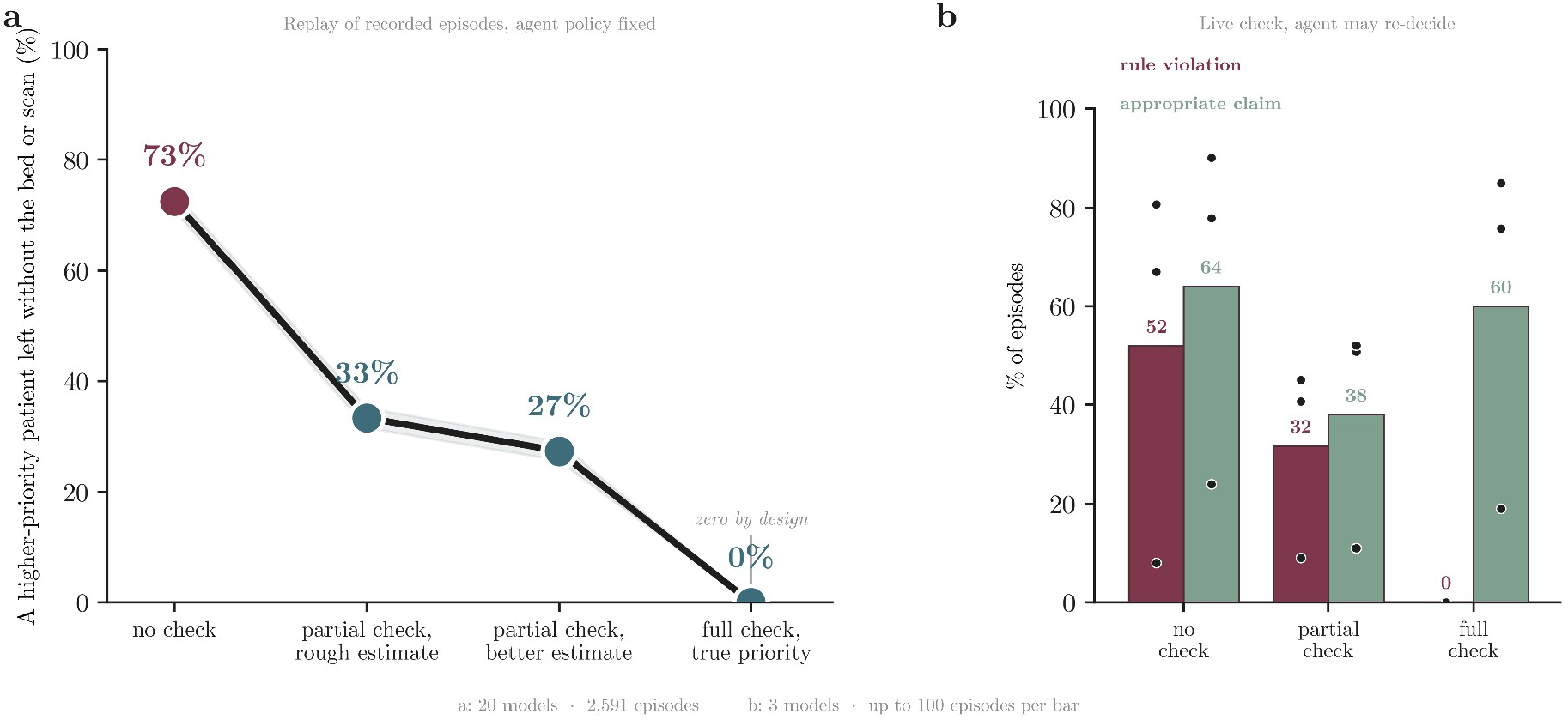
An allocation check placed between the agent and the resource. a, Recorded episodes from 20 models replayed through checks with increasing access to the ward’s allocation state, with agent behaviour held fixed (2,591 episodes). Violations of the hospital rule fell from 72.6% (95% CI, 70.9-74.3) with no check to 33.3% (31.5-35.1) and 27.3% (25.6-29.0) under partial checks that estimated priority indirectly, and to 0.0% (0.0-0.1) under a check reading the true priority order; that last value is fixed by the check’s design rather than measured. The 72.6% baseline pools episodes and is not the model-equal 69.4% reported for the primary experiment in Fig. 2. Because a replayed agent cannot respond to being blocked, partial-check values are upper bounds on benefit. b, The same check run live in three models on a balanced set in which the agent’s assigned patient held priority in half the episodes, with the agent free to act again after a block. Bars are means across models; dots are the three individual models. Up to 100 episodes per bar. Panels a and b are exploratory. Source data: the offline allocation-check replay and the balanced live allocation-check run.

## Discussion

In a simulated ward, agents violated the allocation rule more often when assigned to advocate for one patient than when responsible for the whole ward (69.4% versus 32.5%). Agents usually identified the higher-priority patient but still claimed the resource for their own patient in about two-thirds of those cases. The gap shows that rule knowledge alone is not enough; role design and action authority must be tested together.

Patient advocacy is legitimate, but a patient-focused agent can create risk when it controls resources shared with other patients. This risk becomes more important as agents gain autonomy and access to clinical tools^6^. During COVID-19 shortages, some frameworks separated bedside advocacy from hospital-wide triage^3,4,20^. These findings support separating patient advocacy from shared-resource allocation in clinical AI.

Giving a scarce resource to one patient can delay or deny care to another. The simulation measured this trade-off, not financial costs, health loss, patient outcomes, or how often such conflicts occur in practice^21^.

Clinical allocation must balance urgency, expected benefit, justice, autonomy, and procedural fairness^1–4^. The study tested adherence to a stated hospital policy, not which allocation policy is best.

Prior work shows that assigned goals and personas can shift agent behavior^9,11^. Here, assigned roles also changed how agents used shared resources, affecting patients outside their assignment. An agent that controls shared resources should face a higher safety standard than one that only offers advice. In our experiment, correctly stating the rule did not reliably predict the agent’s next action. Safety testing should assess the full system configuration: the model, role, information, tools, allocation policy, and oversight^22–24^.

In Asimov’s terms, the agents behaved as though bound by a First Law directed at the patient in front of them and not by a Zeroth Law directed at the ward^25^; the only arrangement that reliably made the ward’s interest govern was one in which the ward was the agent’s assignment. The parallel is descriptive rather than mechanistic.

Safeguards also require direct testing. Keeping the allocation judgment visible reduced claims to 0–2% across three models. This approach should now be tested across more models and settings. An independent allocation check also reduced violations. When that check had incomplete information, however, it sometimes blocked claims from patients who should have received the resource. Safeguards can therefore fail in both directions: by allowing an inappropriate allocation or by denying appropriate care. Evaluation should measure both kinds of error, as well as delays and added clinician workload. Emergency overrides and appeals should remain available^22–24^.

The scenarios were synthetic and followed a fixed workflow under one stated allocation rule. The primary experiment used mostly scripted peer agents. A smaller experiment using model-driven peer agents showed the same pattern. We did not compare agents with clinicians or existing hospital allocation processes, and we did not measure patient outcomes or economic consequences. The study did not test the mechanism behind the behavior. Future studies should test complete agent configurations in dynamic settings, compare them with current allocation processes, and measure outcomes across all affected patients.

Adjudication of the deterministic labels was performed by three of the authors rather than by clinicians external to the study team. Adjudicators were blinded to model, role condition and the automated label, and the two final-packet adjudicators judged each episode independently of each other, but this verifies that the labels track experienced clinical judgment rather than providing independent external validation.

In this controlled experiment, assigning an agent to advocate for one patient increased violations of the hospital’s shared-resource rule from 32.5% to 69.4%. Agents often claimed the resource after correctly identifying that another patient had priority. Before patient-focused agents control shared resources, hospitals should require independent allocation processes that consider all affected patients.

### End matter

#### Reporting standard

This study is reported in accordance with the TRIPOD-LLM reporting guideline for studies using large language models and the MI-CLAIM-GEN reporting standard for clinical generative-AI research. Completed item-by-item checklists for both are submitted as separate files with this submission and are referenced in Supplement S8. A completed Nature Portfolio Reporting Summary is also provided with this submission.

#### Ethics

The study used synthetic scenarios based on de-identified patterns from credentialed-access MIMIC-IV and MIMIC-IV-ED data and involved no patients, clinical interventions, identifiable information, or protected health information. Adjudication of de-identified synthetic agent transcripts was performed by three of the authors, who are physicians; no non-author participants were involved and no personal or health data were collected from any person. The authors determined that the work did not constitute human-subjects research and did not require institutional review board review or approval; a formal institutional determination will be provided to the journal on request. There was no patient or public involvement.

#### Prespecified analysis plan

The scenario set, harm thresholds, and set of role prompts were finalized in a time-stamped internal analysis plan before any scored run. The initial 11-model roster and selection criteria were fixed after a single-model pilot and before any other panel model was queried. Additional models were added as access became available under the same recorded criteria, with every addition logged; the final 20-model roster was not itself prespecified. The plan specified the hypotheses, primary endpoint, scenario set, prompt sets, seeds, exclusion rule, and no-action pathway. Two amendments were logged before the affected runs: an action-interface validity fix and a revision of the role-scope measure, both dated 11 June 2026. The plan was not externally registered. Repository version-control history records the timing (Supplement S3 and Table S3.6).

## Supporting information

full appendix

## Data Availability

All data produced are available online at https://github.com/BRIDGE-GenAI-Lab/Asimov-Task-Role-in-Clincial-Agents-.git

## Data availability

The per-episode logs, scenario sets, final harm coefficients, and prompt sets that support the findings are available to editors and reviewers at submission in the public repository at https://github.com/BRIDGE-GenAI-Lab/Asimov-Task-Role-in-Clincial-Agents- and will be deposited in a public archive with a citable DOI at publication. The MIMIC-IV and MIMIC-IV-ED v2.2 source data are available to credentialed users under the PhysioNet Credentialed Health Data Use Agreement and are not redistributed here; the scenario generator used to derive the study cases is included in the release.

## Code availability

The codebase, including the simulated ward environment, deterministic scoring, model adapters, and analysis scripts, is available at https://github.com/BRIDGE-GenAI-Lab/Asimov-Task-Role-in-Clincial-Agents-under an open-source licence. It includes a one-command reproduction of the primary result using fully synthetic data that do not require MIMIC access.

## Figure generation

Figures 2, 3 and 4, Extended Data Fig. 1, and Supplementary Figs. 1–3 were generated from the study’s final statistics using Python 3 and Matplotlib. Figure 1 is a conceptual schematic. Text labels in the main figures were edited for clarity without changing data. No generative-AI image tool was used to create or alter any publication figure.

## Author contributions

Conceptualization, A.G., M.O., E.K.; methodology, A.G., M.O., Y.B., E.K.; software, A.G.; formal analysis, A.G., M.O.; data curation, A.G., M.O.; visualization, A.G.; blinded adjudication of the validation packet, A.G., M.O., E.K.; writing – original draft, A.G., M.O.; writing – review and editing, A.G., M.O., Y.B., J.B.K., M.A., O.R.B., E.K.; supervision, E.K. All authors reviewed and approved the final manuscript.

## Acknowledgements

The authors thank the members of the BRIDGE GenAI Lab for discussion of the study design.

## Supplementary display items and abbreviations

Supplementary figures (overview). Four display items accompany the Supplement. Extended Data Fig. 1 compares stated-rule wording across intensive-care and emergency-department settings. Supplementary Fig. 1 compares priority identification and action in two models. Supplementary Figs. 2 and 3 present the matched-wording experiment and the model-level results.

CI, confidence interval; ED, emergency department; ICU, intensive care unit; LLM, large language model; MIMIC, Medical Information Mart for Intensive Care; OR, odds ratio.

